# Performance of progressive and adaptive COVID-19 exit strategies: a stress test analysis for managing intensive care unit rates

**DOI:** 10.1101/2020.05.16.20102947

**Authors:** Jan-Diederik van Wees, Martijn van der Kuip, Sander Osinga, David van Westerloo, Michael Tanck, Maurice Hanegraaf, Maarten Pluymaekers, Olwijn Leeuwenburgh, Lonneke van Bijsterveldt, Pien Verreijdt, Logan Brunner, Marceline Tutu van Furth

## Abstract

**Background:** In May 2020, many European countries have begun to introduce an exit strategy for the coronavirus disease 2019 (COVID-19) pandemic which involves relaxing social distancing measures. Predictive epidemiological modeling indicates that chances for resurgence are high. However, parametrization of the epidemiological nature of COVID-19 and the effect of relaxing social distancing is not well constrained, resulting in highly uncertain outcomes in view of managing future intensive care unit (ICU) needs.

**Methods and findings:** For performance analysis of exit strategies we developed an open-source ensemble-based Susceptible-Exposed-Infectious-Removed (SEIR) model. It takes into account uncertainties for the COVID-19 parametrization and social distancing measures. The model is calibrated to data of the outbreak and lockdown phase. For the exit phase, the model includes the capability to activate an emergency brake, reinstating lockdown conditions. Alternatively, the model uses an adaptive COVID-19 cruise control (ACCC) capable to retain a targeted ICU level. The model is demonstrated for the Netherlands and we analyzed progressive and adaptive exit strategies through a stress test of managing ICU rates. The progressive strategy reflects the outcome of social and economic pressure to use one-way steering toward progressively relaxing measures at an early stage. It is marked by a high probability for the activation of the emergency brake due to an unsolicited growth of ICU needs in the following months. Alternatively, the two-way steering ACCC can flatten ICU needs in a more gradual way and avoids activation of the emergency brake. It also performs well for seasonal variation in the reproduction number of severe acute respiratory syndrome-coronavirus.

**Conclusions:** The adaptive strategy (ACCC) is favored, as it avoids the use of the emergency brake at the expense of small steps of restrictive measures and allows the exploration of riskier and potentially rewarding measures in the future pathways of the exit strategy.

## INTRODUCTION

According to the World Health Organization (WHO), coronavirus disease 2019 (COVID-19) reached the pandemic phase on 11 March 2020 (1). On 15 May 2020, the severe acute respiratory syndrome-coronavirus (SARS-CoV-2) virus had spread to 213 countries worldwide, leading to 4,586,270 registered infections and 306,063 deceased (2). To control COVID-19, various measures have been undertaken ranging from liberal control in Sweden and test and quarantine policies in South Korea, to complete prolonged lockdowns in Italy and Spain (3). The chosen measures of national policymakers are based on testing capacity, characteristics of specific societies and more importantly on the phase of the outbreak: containment, suppression or mitigation (4). ‘All countries must strike a fine balance between protecting health, preventing economic and social disruption, and respecting human rights’ (1). In the Netherlands, a targeted lockdown based on social distancing, was introduced on 16 March 2020 (5).

As the trend of death rates (Fig. 1) and hospital admissions are slowly showing a flattening curve and intensive care units (ICU) no longer work at or above their capacity, many countries have begun to introduce an exit strategy which involves relaxing social distancing measures. Predictive epidemiological modeling demonstrated that prolonged and/or intermittent periods of social distancing are required to mitigate the possibility of resurgences of infection (6, 7). They have not been calibrated to (near) real-time data and did not consider quantitative measurement and control systems to assess and mitigate unsolicited stress to the health care system. Moreover, these models highlight that parametrization of the epidemiological nature of COVID-19 is not well constrained and highly uncertain (4, 6–8). For this reason, models may be perceived as unreliable in light of all the uncertainties considered, and in turn the lack of reliable models causes a major challenge in robustly designing and testing exit strategies.

**Figure 1.**
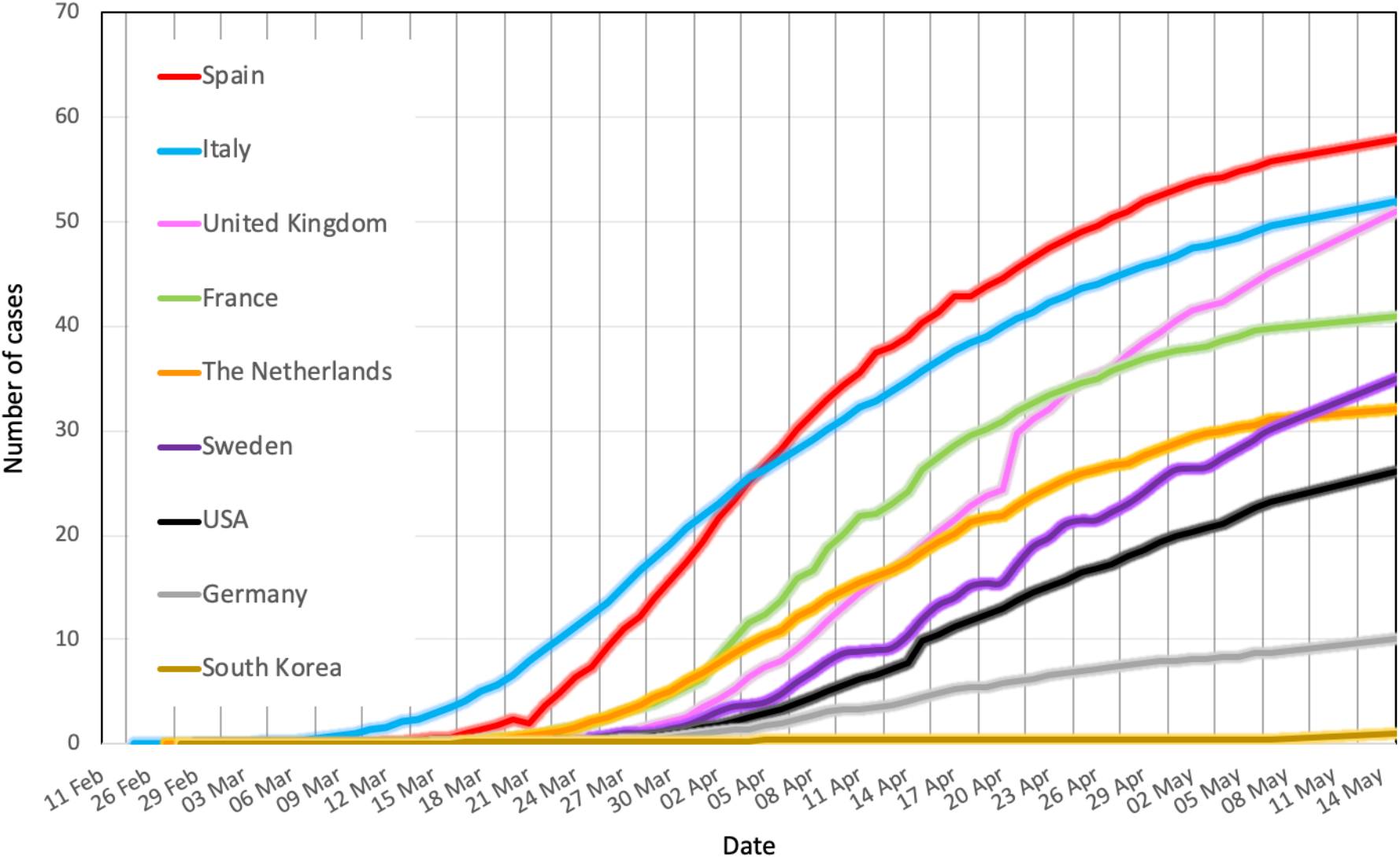
COVID-19 mortality per 100,000 population per country (2) The Netherlands rank intermediate (orange line) between Spain / Italy (high mortality) and Germany / South Korea(low mortality).

In order to cope with large uncertainties for correct parametrization of transmission models and the effect of social distancing measures, we propose a data-driven and holistic approach for calibrating transmission models and the effects of non-pharmaceutical interventions under large uncertainty. To this end, we used a fast, ensemble based Susceptible-Exposed-Infectious-Removed (SEIR) meta-population model explained in methods, which can be calibrated daily to observed data. As a starting point for the exit strategy, the model uses the posterior ensembles and the underlying model and intervention parameters, which have been calibrated to data of the outbreak and lockdown phase (Fig. 2). This provides a robust model basis for a (real-time) analysis and stress test of appropriate exit strategies, and underlying uncertainty. Furthermore, since the model is open-source and can build from public data sources and is not dependent on detailed epidemiological field study results, such as testing or contact-tracing information, it can contribute to the need of the scientific community for accelerated development of open modeling approaches to study exit strategies (9).

**Figure 2.**
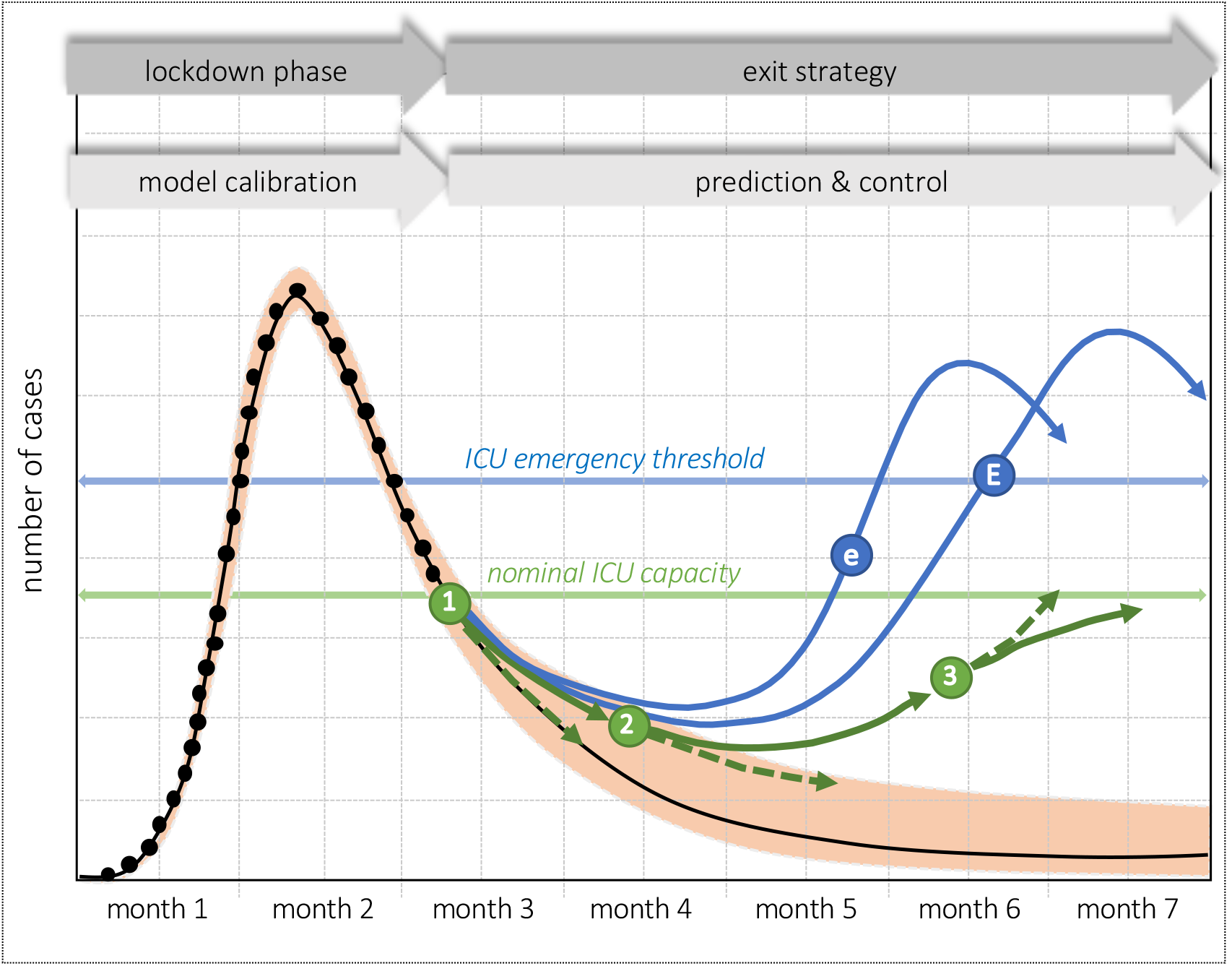
**Schematic diagram of ICU needs, rapidly growing during the initial outbreak and subsequently deflected under lockdown and social distancing conditions**. The exit strategy is aimed at a tradeoff between limited spreading and maximizing relaxation of social distancing. Two exit strategies have been considered: a progressive relaxation (blue arrows), and alternatively the adaptive COVID-19 cruise control (ACCC-green arrows). In the progressive scenario, an emergency brake is activated if ICU needs grow beyond the ICU emergency threshold level **(E)**, or if the daily ICU growth rate exceeds a threshold level **(e)**. The ACCC incorporates a biweekly adjustment of social distancing measures to flatten toward a targeted nominal ICU capacity. Relaxed social distancing steps are taken at evaluation toll gates **(1)** and **(2)**. The relaxing measures need to be adjusted in time to constraining measures to avoid unsolicited growth above nominal ICU capacity. To this end, restrictive measures are taken at toll gate **(3)**. For the ACCC, the dashed and solid arrows denote the default and adjusted ICU trends due to the measures at the tollgates.

Two ‘safe’ exit strategies as outlined in Fig. 2 have been put to a stress test in a case study for the Netherlands. Both aim at maximizing the positive effect on social distancing and reducing risks for the healthcare system at the same time. They consider (step-changed) relaxation in conjunction with possible restrictive measures, in view of managing the risk for exceeding threshold levels for ICU capacity. The two strategies differ fundamentally in ICU objectives and approach. The progressively relaxing and ‘emergency brake’ strategy reflects the outcome of social and economic pressure to use one-way steering toward progressively relaxing and rather early steps and it is marked by a high probability for the need of the use of an emergency brake due to an unsolicited growth of ICU needs in the following months. Alternatively, the Adaptive COVID-19 Cruise Control (ACCC) is based on two-way steering and can flatten ICU needs in a more gradual way to a given nominal capacity. We will show that the ACCC strategy can avoid the need for an emergency brake, and also performs well if the potential effect of seasonal variation in the reproduction number of SARS-CoV-2 is taken into account (7).

## METHODS

For the modeling, we used an ensemble-based SEIR metapopulation model to simulate the epidemics (8, 10), which incorporates the effect of measures to reduce community spreading (4, 8). The model is largely based on van Wees et al., 2020 (11), and has been expanded in this study to incorporate a more realistic flow process for hospitalization and ICU (Fig. 3).

**Figure 3.**
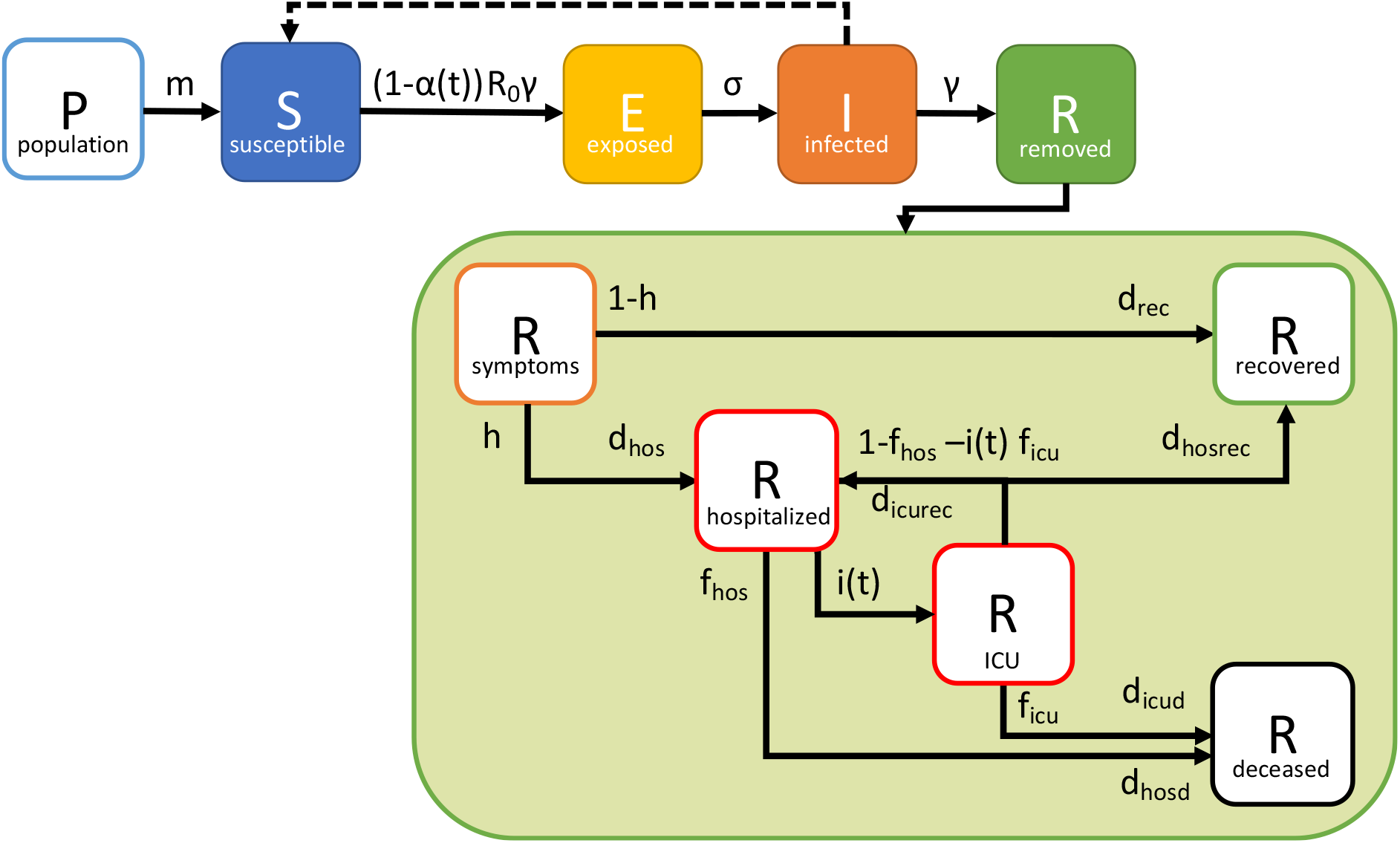
**Schematic diagram of the SEIR model, extended for R with sub-compartments for the flow of hospitalized patients (fraction *h*), and subsequently ICU patients (fraction *i*(*t*))**. The recovery time from mild symptoms, delay and treatment times for hospitalization, and ICU are marked by *d_hos_, d_rec_, d_hosrec_* and *d_icurec_*, respectively. The CFR and associated treatment periods for hospitalized and ICU fatal patients are marked by *f_hos_, d_hosd_* and *f_icu_, d_icud_*, respectively. Recovered ICU patients first flow back to hospital and are assumed to take *d_hosrec_* for full recovery. Each of the parameters can be marked by a priori constants or distributions, which can be adjusted in the data calibration. Table 1 lists the adopted parameter values including the prior and posterior parameters used for the case study of the Netherlands. Some of the treatment times are marked by a logistic spread represented by a Gamma distribution (see Table 1).

The SEIR model stands for the following: Susceptible: the susceptible part of the population; Exposed: the part of the population that carries the virus; Infected: patients showing the first mild symptoms; Removed: patients that have recovered or died. Mathematically, the SEIR compartments are modeled as fractions of the population, initially susceptible to the virus. Subsequently, we solve the following differential equations starting from initial values for *S* = 1–1/*N, E* = 1/*N* and setting *I* and *R* to 0. *N* is ratio of population size and exposed persons introduced at the start of the simulation. The initial day of introduction of the virus is 15 days prior to the first registered case, and the density of the virus per *N* is varied to obtain a first order fit with the observed number of registered infections, hospitalized, death. The differential model is formulated as follows:

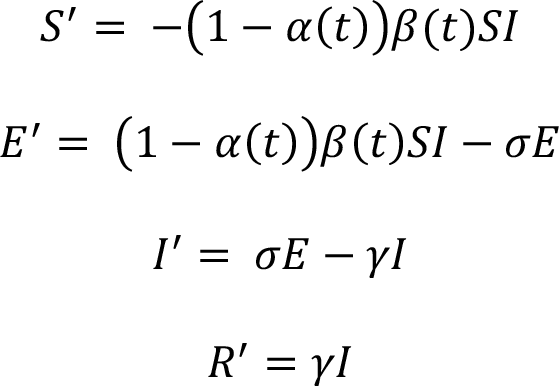

The *R* compartment is extended with the flow scheme for the hospitalization and ICU treatment process as depicted in Fig. 3. For the incorporation of the logistic spread of patients, the model adopts successive convolutions of the *R* prediction, with a Gamma distribution for delay, recovery and treatment times *d_hos_, d_hosrec_, d_hosd_, d_icurec_* and *d_icud_* (Fig. 3, Table 1). The simulation incorporates the strength of social distancing through the parameter *α(t)*: *α(t)* is 0 when no measures are in place and 1 when social distancing would prevent any transmission (8). The ensemble-based workflow intrinsically allows for the use of large uncertainties underlying the relaxing and restrictive steps in social distancing and can therefore be used effectively for stress tests of exit scenarios. Our model is strongly data-driven and calibrates prior estimates for a number of model parameters (Table 1), including the reproduction number, strength of social distancing, and some of the treatment times for hospitalization and ICU from the observed data. For data assimilation of all parameters in conjunction with calibration of *α(t)*, we use the ensemble smoother with multiple iterations which is a computationally efficient method for ensembles of non-linear forward models (12). The data assimilation is performed by matching modeled hospitalization and ICU usage to reported usage. We adopted ensembles of 500 realizations and a value of 70 and 20 for the error in standard deviation in reported bed count for hospitalization and ICU, respectively.

**Table 1.**
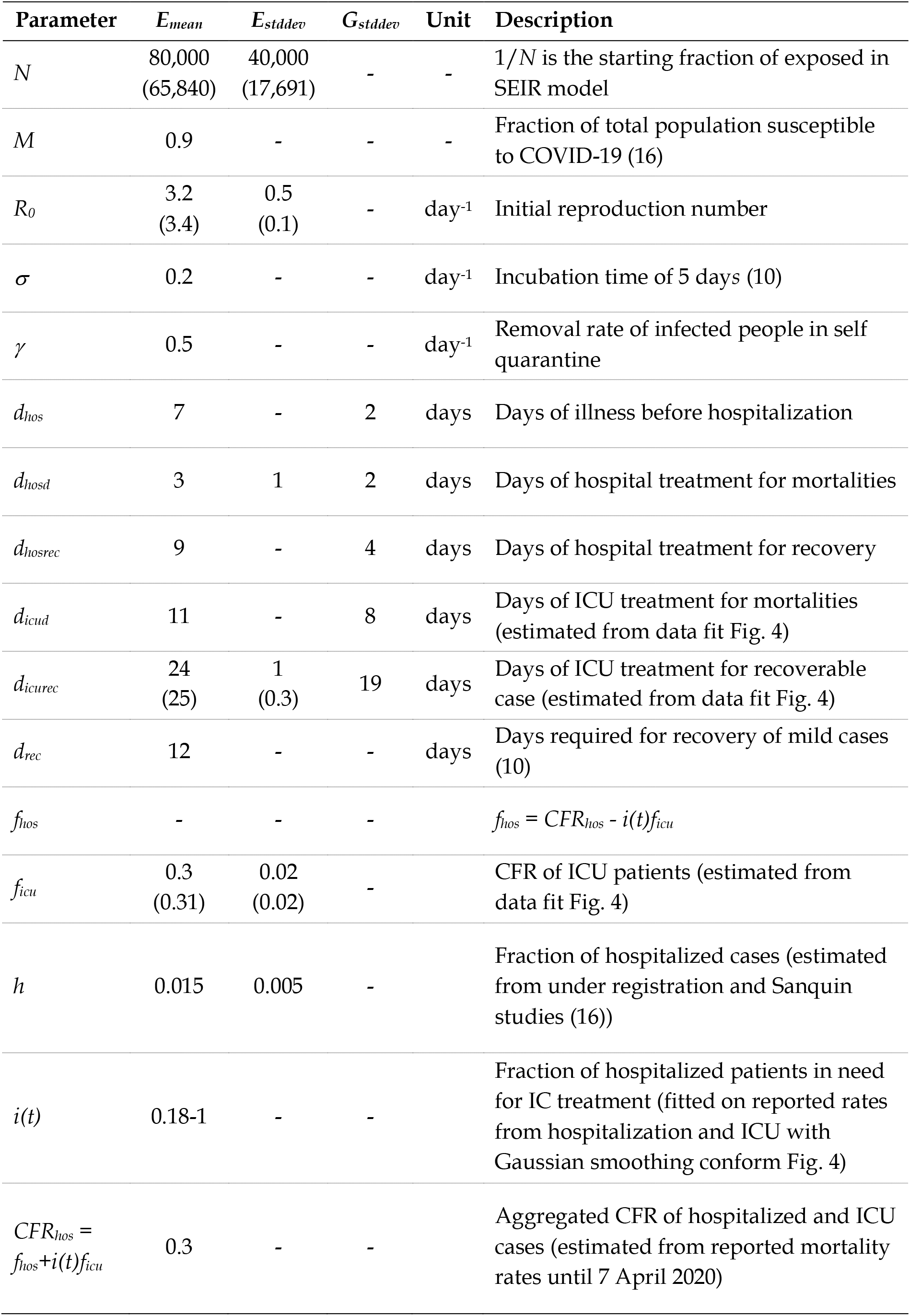
**Parameters for the SEIR model**. The mean and standard deviation of prior and posterior (in parentheses) normal distributions of the model parameters. For each ensemble member, the patient (treatment) times *d_hos_, d_hosrec_, d_hosd_, d_icurec_* and *d_icud_* are represented with a Gamma distribution with mean sampled from *N (E_mean_, E_stddev_*) and standard deviation of *G_stddev_*.

| Parameter | $E_{mean}$ | $E_{stddev}$ | $G_{stddev}$ | Unit | Description |
| --- | --- | --- | --- | --- | --- |
| $N$ | 80,000<br>(65,840) | 40,000<br>(17,691) | - | - | $1/N$ is the starting fraction of exposed in SEIR model |
| $M$ | 0.9 | - | - | - | Fraction of total population susceptible to COVID-19 (16) |
| $R_0$ | 3.2<br>(3.4) | 0.5<br>(0.1) | - | day<sup>-1</sup> | Initial reproduction number |
| $\sigma$ | 0.2 | - | - | day<sup>-1</sup> | Incubation time of 5 days (10) |
| $\gamma$ | 0.5 | - | - | day<sup>-1</sup> | Removal rate of infected people in self quarantine |
| $d_{hos}$ | 7 | - | 2 | days | Days of illness before hospitalization |
| $d_{hosd}$ | 3 | 1 | 2 | days | Days of hospital treatment for mortalities |
| $d_{hosrec}$ | 9 | - | 4 | days | Days of hospital treatment for recovery |
| $d_{icud}$ | 11 | - | 8 | days | Days of ICU treatment for mortalities (estimated from data fit Fig. 4) |
| $d_{icurec}$ | 24<br>(25) | 1<br>(0.3) | 19 | days | Days of ICU treatment for recoverable case (estimated from data fit Fig. 4) |
| $d_{rec}$ | 12 | - | - | days | Days required for recovery of mild cases (10) |
| $f_{hos}$ | - | - | - | | $f_{hos} = CFR_{hos} - i(t)f_{icu}$ |
| $f_{icu}$ | 0.3<br>(0.31) | 0.02<br>(0.02) | - | | CFR of ICU patients (estimated from data fit Fig. 4) |
| $h$ | 0.015 | 0.005 | - | | Fraction of hospitalized cases (estimated from under registration and Sanquin studies (16)) |
| $i(t)$ | 0.18-1 | - | - | | Fraction of hospitalized patients in need for IC treatment (fitted on reported rates from hospitalization and ICU with Gaussian smoothing conform Fig. 4) |
| $CFR_{hos} = f_{hos} + i(t)f_{icu}$ | 0.3 | - | - | | Aggregated CFR of hospitalized and ICU cases (estimated from reported mortality rates until 7 April 2020) |

Once the model has been calibrated to the data of the outbreak and lockdown phase, in the exit strategies the relaxing or restrictive measures for social distancing are adopted by a step-wise change of the posterior *α(t)* values, such that *α(t>t_step_) = α(t)+ Δα(t)* at given times or at specific time intervals *t_step_*. In the model, *Δα(t)* = *N*(*m*_step_, *s*_step_) of each step is normally distributedwith *m*_step_ = mean and *s*_step_ = standard deviation. For each member in the ensemble, a sample is taken for *Δα(t)*. Based on the updated *α(t)*, the model is rerun. The resulting ICU occupancy is monitored daily in the model. If the ICU occupancy or the ICU growth rate exceeds a given level, the emergency brake is activated. The emergency brake reinstates lockdown conditions by adopting *α(t>t_emergencybrake_)* = *N*(*m_lockdown_,s_lockdown_*), where *m_lockdown_* and *s_lockdown_* are based on the posterior distribution of *α(t*) in the data calibration of the lockdown phase. In the aftermath of the emergency brake, the renewed lockdown conditions can be followed by renewed steps for relaxation of social distancing, once the resulting ICU occupancy drops to acceptable levels. In the progressive exit strategy only user-defined relaxing measures (*m_step_*<0) are considered for social distancing.

In the adaptive exit strategy, we consider the ACCC to automatically steer both relaxing (*m_step_*<0) and restrictive step-changes (*m_step_*>0) toward reaching and retaining a sustainable level of nominal ICU capacity and to avoid activation of the emergency brake and renewed lockdown conditions. To this end, the need for a step-change and its sign is monitored at intervals of two weeks, taking the actual ICU occupancy, ICU growth rate and change in growth rate of the past two weeks of the ensemble member, and then using an extrapolated value from a second-order Taylor approximation that looks one week ahead. If the extrapolated growth rate (and value) is moving away from the targeted nominal capacity, a relaxing (when below target) or restrictive (when above target) step is adopted. A constrictive step is taken if the change in growth rate is accelerating toward the target. For the time following the first relaxation steps after the lock down, this effectively means that steps are taken to slow down the growth rate as soon as the look ahead signals a rise in ICU occupancy.

## RESULTS

### The Netherlands’ outbreak and lockdown phase

With a population density of ∼1,300/mi.^2^, the Netherlands (∼17.1 million citizens) ranks number 32 worldwide in population density (in perspective New York state: ∼19.5 million citizens and ∼420/mi.^2^ and USA overall density of ∼95/mi.^2^) (13). In the era before COVID-19, the ICU capacity was 1,150 and occupied for ∼75%. In the Netherlands, the first patient with COVID-19 was diagnosed on 27 February 2020. At the peak of the outbreak on 2 April 2020, 1,332 patients with COVID-19 were admitted at an ICU. The current maximum national capacity is 1,800 ICU including non-COVID-19 care (14) In the remainder of this paper we refer to COVID-19 ICU capacity only.

The daily admittance of ICU patients closely followed the trend of hospitalized patients (Fig. 4). Markedly, the fraction of hospitalized patients that required an ICU shows a gradual decreasing trend toward a relatively constant of 20% from the end of March onwards. The case fatality of ICU patients is 30% (14) and the overall mortality of COVID-19 patients (5,643 on 14 May 2020) in the Netherlands contributes to ~2% of the global death toll of the pandemic (Fig. 1) (2). The cumulative ICU mortality is approximately 10% of the registered COVID-19 death toll in the Netherlands. This can be well explained by the fact that approximately 80% of ICU patients are below the age of 70 (14), whereas 90% of the total mortality is in the age groups of 70 and older (15). Also, hospitalized patients are skewed toward younger patients, with approximately 50% below the age of 70 (16). These numbers highlight that hospital and ICU admittance in the Netherlands is directed towards younger age groups compared to earlier studies assessing hospitalization and ICU needs (4). This admittance practice significantly reduced pressure on the health care system.

**Figure 4.**
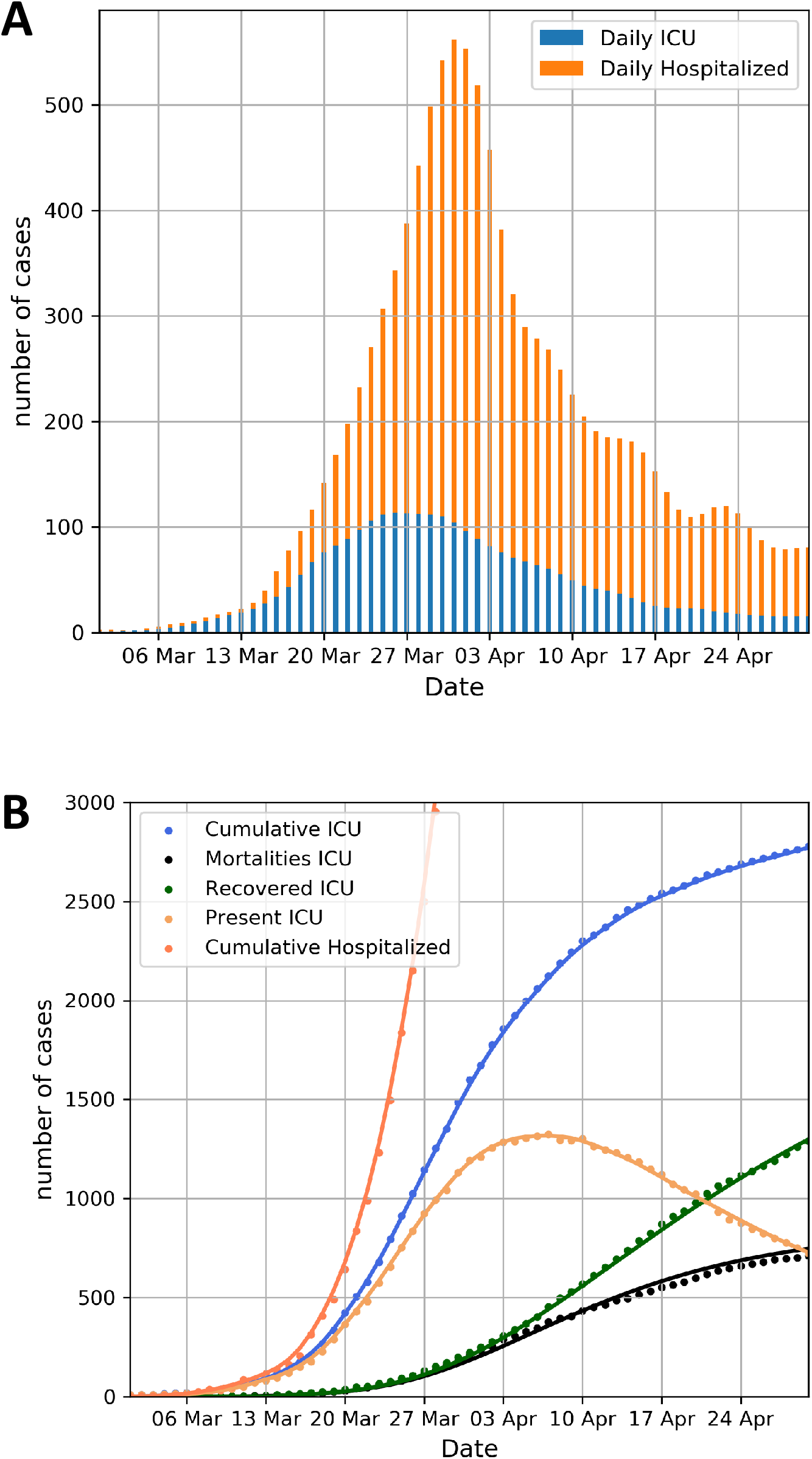
**The Netherlands’ COVID-19 ICU characteristics (period of March and April 2020). (A)** Daily in-hospital and ICU admissions adjusted with a Gaussian smoother with a standard deviation of 1.5 days. **(B)** Cumulative counts of hospitalized and ICU admissions (actual data and smoothed lines). The daily ICU occupancy, cumulative recoveries and mortality (14) have been forecasted from the daily ICU admission data with Gamma distributions for ICU treatment times which differ for mortality and recovery (see Table 1). The estimated ICU mortality rate is 30%.

The forecast presented here, has been based on the data until 30 April 2020, calibrating the prior model parameters to the observed data on hospitalized and ICU patients, as well as mortality prior to 7 April 2020. The forecast includes 500 ensemble members, and 8 iterations were used in the ensemble smoother for the calibration to the data. The time-invariant prior and resulting posterior model parameters are listed in Table 1. In the model prior *α(t)* values, have been chosen based on expert judgement and are in the range of values used in literature (8), based on the logic of contact reduction and social distancing (4, 8). They have been introduced on the dates of government measures (11), with a time-interval of 5–10 days and adopted a relatively large a-priori uncertainty for the social distancing strength *α(t)* with a standard deviation of 10–30% of its estimated value (11). The posterior time-variant strength of the effect of social distancing, reducing the transmission strength by 1-*α(t)*, and the predicted model results for infected, hospitalized and ICU patients and mortality are shown in Fig. 5A-E. The social distancing strength (Fig. 5A) have been adjusted in the ensemble smoother to fit data on hospitalized (Fig. 5C) and ICU occupancy (Fig. 5D). The model is very well capable of reproducing the observed hospitalization (Fig. 5C) and ICU rates (Fig. 5D) and demonstrates the effectiveness of the social distancing in terms of reducing the hospitalization inflow, resulting in a peak ICU occupancy in early April 2020 followed by a downward trend to manageable levels on 30 April 2020 close to an occupancy of 700 ICU patients. It is expected that ICU usage will further reduce well below 600 on 11 May 2020, the date the Dutch government has been planning to relax the lockdown conditions. The posterior 1-*α(t)* evolution is marked by a rapid stepwise decrease in March 2020, in line with the government restrictions. The confidence bandwidth of the 1-*α(t)* values shown in Fig. 5A is consistent with the data and the prior model parameter uncertainty and possible combinations in temporal change of *α(t)* resulting from data assimilation process. After 7 April 2020 in the model, the *α(t)* has been considered invariable through time, resulting in a relatively narrow bandwidth of future forecasts for ICU needs.

**Figure 5.**
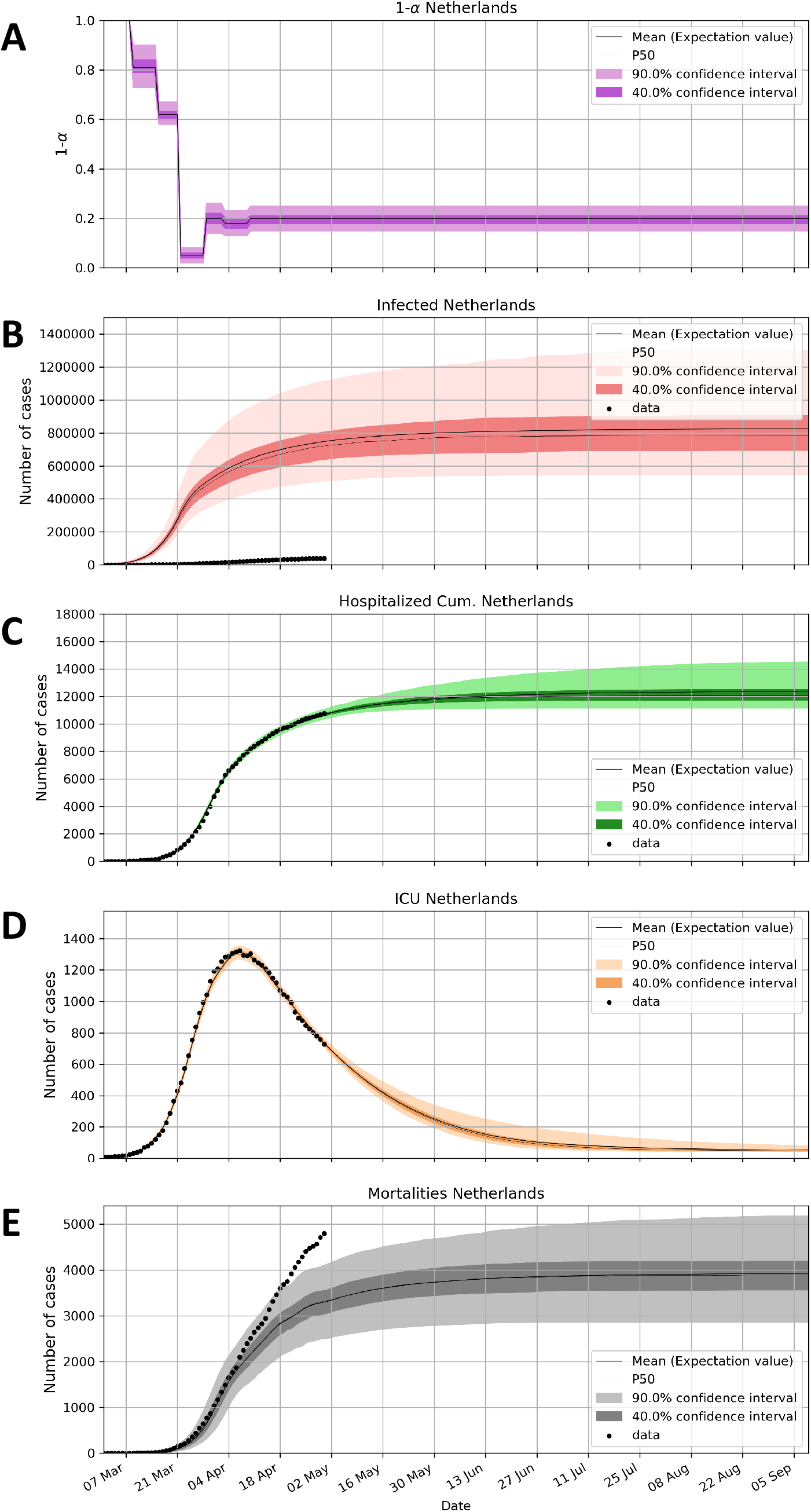
**Calibrated ensemble forecast based on the Netherlands’ data up until the end of April 2020. (A)** relative strength of transmission, due to social distancing measures, relative to the start (1.0) of the outbreak, **(B)** cumulative infected people, **(C)** cumulative hospitalized, **(D)** ICU occupancy, and **(E)** mortality. See text for explanation.

The mortality beyond 7 April 2020 is significantly higher than expected from the hospitalization flow model. One possible explanation for this strong deviation, is that a growing share of the registered deceased patients are related to care centers dedicated to elderly people (Fig. 5E). Here COVID-19 has been marked by more active spreading than the national trend in the last three weeks of April 2020. This may be related to a lack of testing and protective measures for care-giving personnel in that setting of elderly care. It should also be noted that overall many more casualties are related to COVID-19 than tested. It is estimated from nationally recorded death rates that the real death toll of COVID-19 may be twice as high as the registered rates indicate (15). The same applies to estimates of the fraction of the Dutch population which may have been infected. Based on blood samples from Sanquin, the Dutch national blood bank, up to 3.6% of the Dutch population may have been infected since the outbreak (17). For this reason, we assumed in the prior of the model prior parameters that a relatively low fraction (1.5%) of those infected people gets hospitalized. Consequently, the predicted number of cumulative infected is significantly higher in the model than actual confirmed cases (Fig. 5B). The SEIR model takes into account the effects of gradual build-up of immunity through the gradual reduction of *S* in the mathematical formula (see methods). Such a reduction can potentially contribute to social relaxation and therefore the estimation of the ratio of infected and hospitalized patients is important. However, with the adopted parameters, the effect of immunity build-up is rather low and not of significant influence for the presented results in this paper.

### Progressively relaxing exit strategy with emergency brake

For the progressively relaxing strategy we considered six scenarios (Fig. 6A-F), in order to highlight key aspects of this approach and the effects of the emergency brake. The first scenario is marked by a relatively large step-change, reducing on 11 May 2020 the social distancing strength *α(t)* from approximately 0.8 to 0.6 with *Δα(t)* = *N*(−0.2,0.05). In absence of renewed mitigation measures, the ICU needs would grow unboundedly within a couple of months as displayed in Fig. 6A.

**Figure 6.**
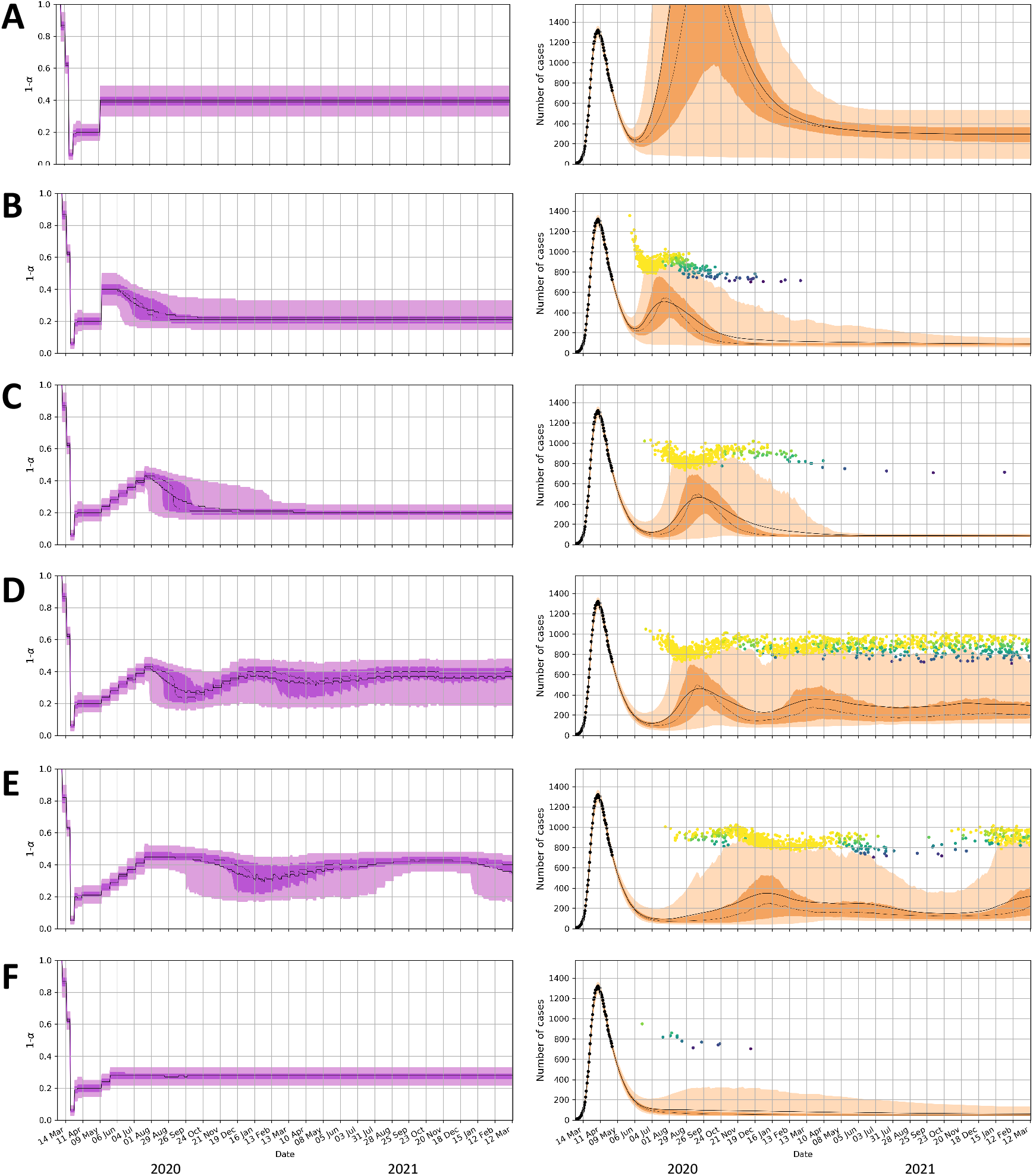
**Progressively relaxing exit strategy. (A)** Large step-change with no emergency brake. **(B)** Same as panel A but with emergency brake **(C)** Small step-changes with emergency brake. **(D)** same as panel C with restart of relaxation when ICU < 500. **(E)** same as panel D with marked by seasonal fluctuation of *R_0_*, with a cosine function ranging from *R_0_*∼3.4 on 1 March 2020 to *R_0_*∼2.4 on 1 September 2020. **(F)** same as panel C with two progressively relaxing steps. Figure conventions same as **Fig. 3**. The dots in the right-hand panels depict the height and time of the peak ICU reached after each instance the emergency brake was activated in the ensemble members. The color of the dots corresponds to the daily ICU growth at the time of triggering the emergency brake (see **Fig. 7**).

In the second scenario (Fig. 6B), the first scenario is complemented with an emergency brake. This emergency brake is triggered either by reaching 700 ICU occupancy or if the daily growth rate in ICU occupancy exceeds 20 (depicted with the colored dots in the right panels of Fig. 6). The underlying full ensemble results of this scenario are also shown in Fig. 7 to highlight the functioning of the emergency brake. The brake reinstates the proven social distancing measure of the lockdown phase with *α(t)* = *N*(0.8,0.05). Effectively, this results in peak ICU values which can reach up to 700–1,400 admissions. Most of the peak values in the ensemble are in the range of 800–1,000 ICU beds, highlighting the long delay between the activation of the brake and its effect in the ICU occupancy. Due to this delay the peak ICU value is expected to show a correlation with the growth rate of ICU occupancy at the time the emergency brake was activated. This can indeed be observed in the ensemble results in Fig. 7

**Figure 7.**
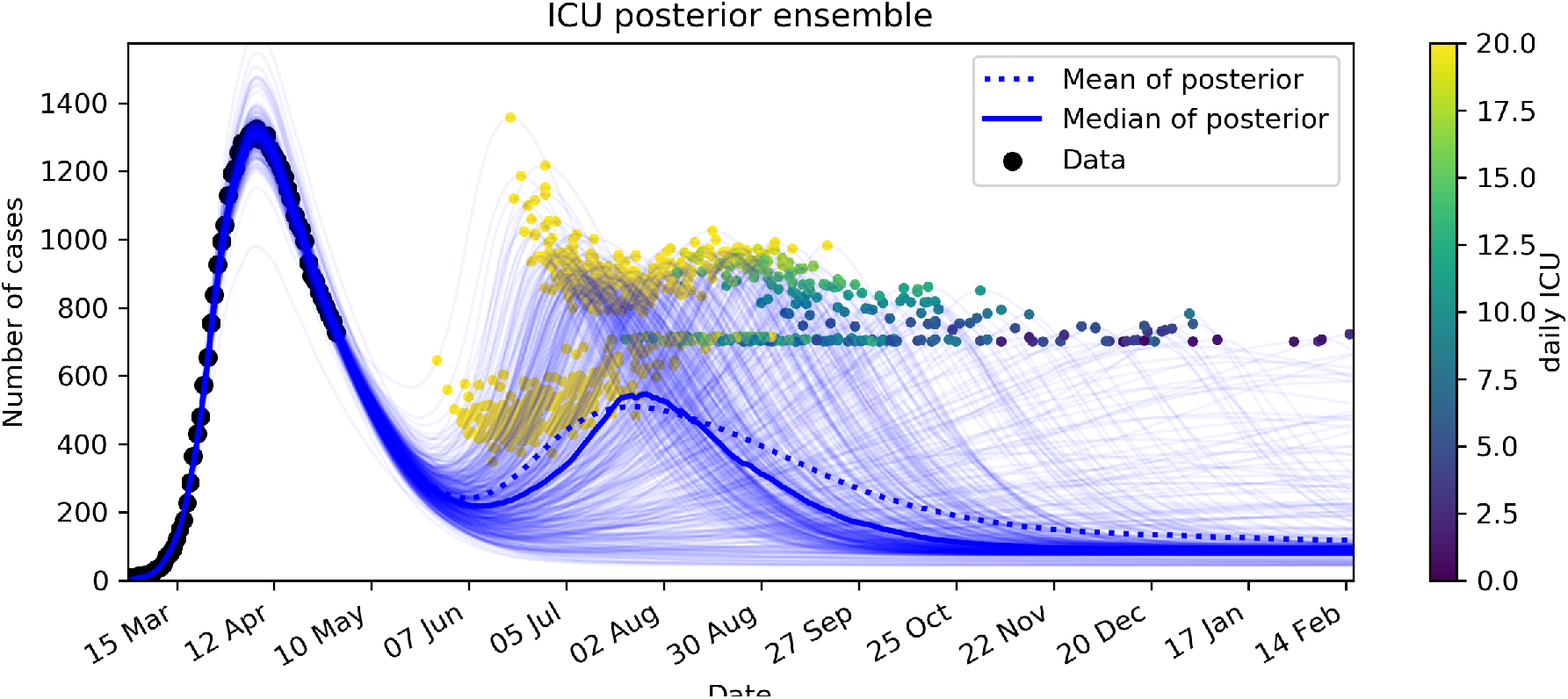
Emergency Brake. Ensemble members (500) for the scenario shown in **Fig. 6B**. Two dots are shown for each member of the ensemble: one at the day and ICU level the emergency brake is triggered and one at the day the peak ICU is reached. The dots are colored according the daily ICU growth rate at the moment of triggering of the emergency brake. Yellow dots represent activation of the emergency brake by daily ICU growth rate>20. Others have been triggered by ICU>700.

In the third scenario (Fig. 6C), the step-change exit strategy adopts the relaxation in five smaller step-changes with *Δα(t)* = *N*(−0.04,0.01), in intervals of three weeks’ time, instead of a single step-change of –0.2. The gradual relaxation results in a more gradual increase in daily ICU needs for the entire ensemble when compared to the single step-change. Consequently, the emergency brake results in a lower peak of ICU admissions, marked by most of the predicted peak values between 700 and 900, and no outliers higher than 1,000.

In the fourth scenario (Fig. 6D), we extend the third scenario with taking progressively relaxing social distancing steps in the aftermath of the emergency brake. The initiation of renewed progressive steps follows after the ICU drops below 500. The relaxing steps can in turn reactivate the emergency brake and consequently a cyclic pattern occurs.

In the fifth scenario (Fig. 6E), we extend the previous scenario with seasonal variation in the reproduction number of SARS-CoV-2 (7) and evaluate its effect on waxing and waning of social relaxation. For the seasonal variation, we adopt a maximum *R_0_* at the start of the model on 1 March 2020 (*R0* ∼3.4), and a minimum at 70% on 1 September 2020 (*R_0_* ∼2.4), varied with a cosine function.

Finally, the last scenario (Fig. 6F) takes a more conservative, progressively relaxing approach in which the number of relaxing steps is limited to 2, and on average *α(t)> (1–1/R_0_*), resulting in a continuous down trend in ICU. In this scenario the emergency brake is only triggered at the start, in a very limited number of members of the posterior. In the remainder of the two years after the two progressive steps no emergency brake is used.

In summary, for the progressively relaxing exit scenario, a large step-change results in a relatively early and possibly high peak, whereas if the change is implemented in more gradual, smaller step-changes, the peak in ICU admissions is delayed and lowered. In both scenarios, the emergency brake is capable of limiting forecasted ICU peaks in bounds of the

1,800-peak capacity in the Netherlands, but any scenario of progressively relaxing steps is at some point likely to result in acceleration of growth and the triggering of the emergency brake. Therefore, progressive step-change scenarios inevitably result in a high probability of a resurgence of infected cases, requiring the need for mitigation measures within the first months up until the first year after the relaxing of social distancing. 1-*α(t)* causes the daily infections and hospitalized cases to rise. Such conditions can also be further amplified in case unsolicited growth is caused by seasonal variation of the reproduction number (7).

### The adaptive COVID-19 cruise control exit strategy

As an alternative to the progressively relaxing exit strategy, we consider the ACCC to steer the step-changes toward reaching and retaining a sustainable level of nominal ICU capacity. For the ACCC’s target ICU nominal capacity, we consider two scenarios: 200 and 400 ICU patients, respectively. We adopt step changes of the same size as the progressive exit strategy *Δα(t)* = *N*(±0.04,0.01). Other settings are the same as in the previous scenarios, except for the emergency brake for the high scenario, where the emergency threshold level for ICU and daily ICU growth have been raised to 1,400 and 40, respectively. The ACCC (Fig. 8A and Fig. 8B) is well capable in steering toward the ICU nominal target levels without activating the emergency brake. The gain in relaxation of social distancing in the high scenario is limited, resulting in *α(t)* which on average is only a fraction lower (∼0.03) than the low scenario, but grows to ∼0.05 in the last year due to slightly faster build-up of group immunity. From the minor gain in social relaxation, it could be argued that the low scenario should be favored, considering the more positive health effects.

**Figure 8.**
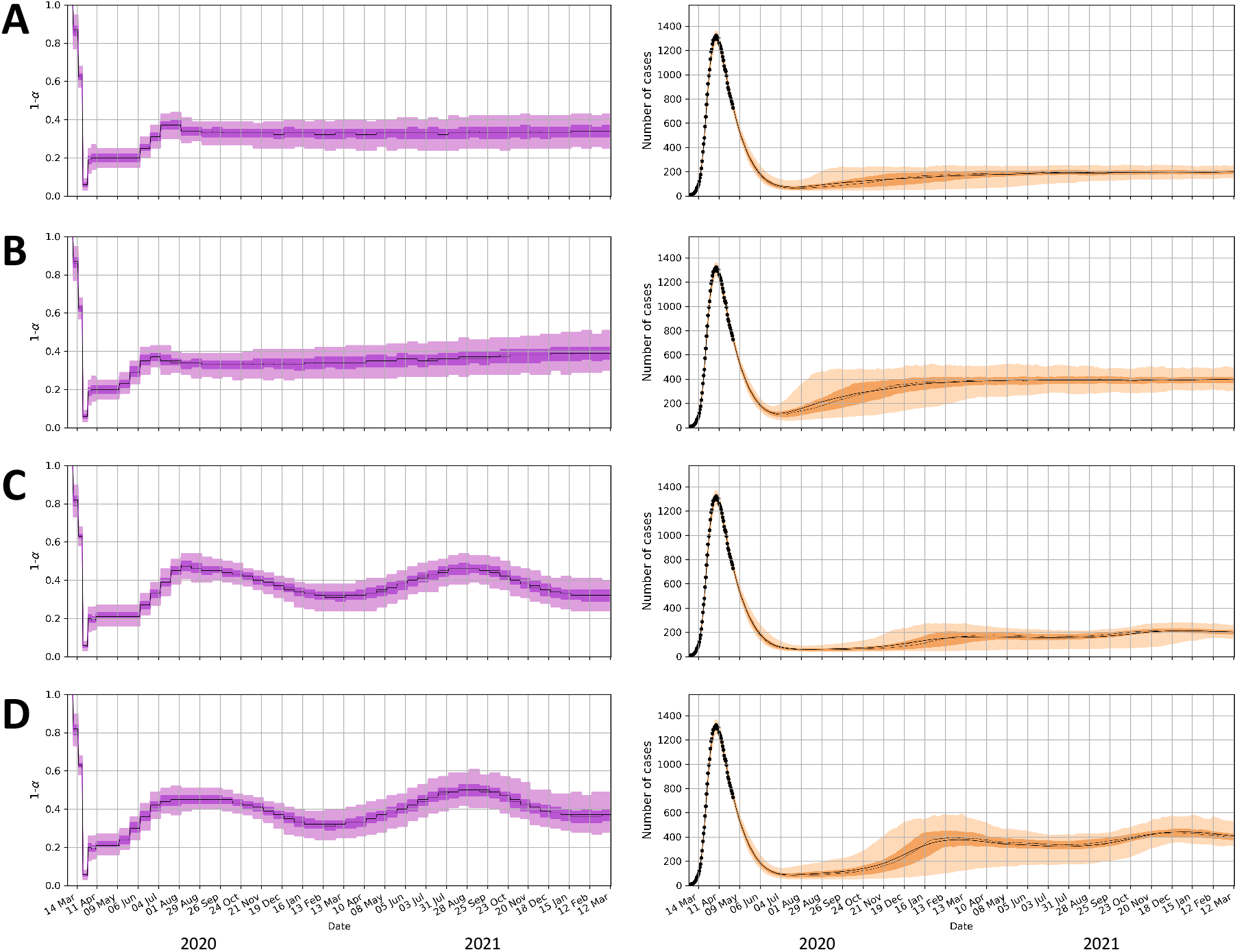
Adaptive COVID-19 cruise control (ACCC) exit strategy for two ICU nominal capacity scenarios. (A, C) ICU capacity of 200 and (B, D) ICU capacity of 400. The panels (C) and (D) are marked by seasonal fluctuation of *R*_0_, with a cosine function ranging from *R*_0_∽3.4 on 1 March 2020 to *R*_0_∽2.4 on 1 September 2020.

Within an alternative scenario, we consider seasonal variation in the reproduction as for the progressive relaxation scenarios (conform Fig. 6E). The results are shown for a period of two years in Fig. 8C and Fig. 8D, causes an appropriate adaptive response of the ACCC, marked by significant seasonal variation in *α(t)*, while maintaining target ICU levels.

In conclusion, the ACCC strategy can effectively steer toward the targeted nominal ICU rates, and the adaptive capability allows it to closely follow seasonal variation in *R_0_*. In both the low and high scenarios, the emergency brake is never activated proving ICU needs remain below 700 and 1,400 ICU respectively at all times for all ensemble runs.

## DISCUSSION

We analyzed two dissimilar ways to exit the lockdown that most countries worldwide are enduring. A progressively relaxing strategy can initially lead to a significant reduction of distancing measures but is likely related to a high peak resurgence of COVID-19 cases for which inevitably an emergency brake will be needed. Alternatively, our ACCC approach leads to small incremental, and constantly evaluated steps in reduction of restrictions and restraining measures if necessary. The ACCC leads to a more gradual and sustainable release, as it does not need to activate the emergency brake. For this reason, we prefer the ACCC approach. Both strategies effectively restrict the number of ICU admissions, however both also indicate that to restrict the number of ICU admissions during an exit strategy the level of safe release of social distancing measures will unfortunately be rather small. In our case study of the Netherlands, increasing total ICU capacity in the ACCC only minimally increases the level of potential release of measures.

A key assumption in both strategies is that we can take limited and controlled step-change sizes. However, in practice, the uncertainty of the effect of each proposed step could be very high. During the exit strategy, assessing and limiting the uncertainty of the effects of each step change is critically important to be able to increase the reliability of prediction and guide the steering of exit strategies. Many proposals for exit scenarios consider further differentiation into age, social, and risk-prone groups, for example, adding to complexity and uncertainty. Evidently, research into transmission characteristics considering different measures can significantly help to reduce uncertainty. However, such an approach can be slow and cumbersome, and will inevitably funnel the choice of controllable steps toward those which are proven, can be tested and monitored and/or are marked by little uncertainty. This can lead to restraints in embracing progressively relaxing steps with uncertain outcomes. The ACCC approach can open pathways toward taking limited risks to empirically explore new avenues in social distancing, since we can timely counteract negative outcomes of relaxing steps with follow-up restrictive steps. The explorative approach can be further enhanced through appropriate diversification of measures at risk, using financial methods such as modern portfolio theory (18) and considering portfolios of uncorrelated measures (e.g. for subpopulations, sectors, age groups, etc.). This will not only help reducing the intrinsic uncertainty of step-changes to much lower values, but at the same time it can accelerate the learning path of the effect of the many different measures considered. In addition, the ACCC can be improved by replacing the rather simple look ahead function by more advanced machine learning and ensemble-based forecasting techniques.

The presented ensemble-based, data-driven and holistic model workflow in this study is fully open source and can promote analysis and further development of exit strategies in other countries, with alternative demographic and transmission characteristics, different social structures, hospital and ICU admittance practices, and a different history of outbreak and lockdown management. Data needed are limited, requiring most importantly hospitalization and ICU admittance, mortality and recovery numbers, and information on past government measures. The emergency brake threshold levels for ICU occupancy and daily ICU growth rate need to be adapted and are recommended to be approximately 40–70% and 0.5–2% of maximum ICU capacity respectively, depending on available lockdown measures and the considered exit strategy.

## Data Availability

We collected the data from publicly available data sources (ref 2, 14, 16 in main text). The modeling code and data used for our analysis is available under (ref 19: github.com/TNO/Covid-SEIR).

https://www.worldometers.info/coronavirus/

https://www.stichting-nice.nl

https://www.rivm.nl/documenten/epidemiologische-situatie-covid-19-in-nederland-6-mei-2020

https://github.com/TNO/Covid-SEIR

## Performance of progressive and adaptive COVID-19 exit strategies: a stress test analysis for managing intensive care unit rates

Jan-Diederik van Wees, Martijn van der Kuip, Sander Osinga, David van Westerloo, Michael Tanck, Maurice Hanegraaf, Maarten Pluymaekers, Olwijn Leeuwenburgh, Lonneke van Bijsterveldt, Pien Verreijdt, Logan Brunner, Marceline Tutu van Furth

## Acknowledgments

We thank Anke Zindler for helpful comments.

## Author contributions

Conceptualization: J-D.v.W., S.O., M.P., O.L., L.v.B., L.B. Data curation: J-D.v.W., S.O., M.P., O.L., L.v.B., L.B., M.T. Investigation: J-D.v.W., S.O., M.P., O.L., L.v.B., L.B. Methodology: J-D.v.W., S.O., M.P., O.L., L.v.B., L.B. Visualization: J-D.v.W., M.v.d.K., PV. Software: J-D.v.W. S.O., M.P., O.L., L.v.B., L.B. Supervision: M.H. Writing, original draft: J-D.v.W., M.v.d.K., M.T.v.F. Writing, review, and editing: all authors.

## Data and materials availability

We collected the data from publicly available data sources (2, 14, 16). The modeling code and data used for our analysis is available under (*19*).

